# Genome-wide association study identifies five risk loci for pernicious anemia and implicates the role of HLA-DR15 haplotype

**DOI:** 10.1101/2020.10.13.20211912

**Authors:** Triin Laisk, Maarja Lepamets, Reedik Mägi

**Author notes:** corresponding author T. Laisk.

## Abstract

Pernicious anemia is a rare condition characterized by vitamin B12 deficiency anemia due to lack of intrinsic factor, often caused by autoimmune gastritis. Patients with pernicious anemia have a higher incidence of other autoimmune disorders, such as type 1 diabetes, vitiligo and autoimmune thyroid issues. Therefore, the disease has a clear autoimmune basis, although the genetic susceptibility factors have thus far remained poorly studied. We conducted a genome-wide association study meta-analysis in 2,166 cases and 659,516 European controls from population-based biobanks and identified genome-wide significant signals in or near the *PTPN22 (*rs6679677, p=1.91 ⨯ 10^−24^, OR=1.63*), PNPT1* (rs12616502, p=3.14 ⨯ 10^−8^, OR=1.70), *HLA-DQB1* (rs28414666, p=1.40 ⨯ 10^−16^, OR=1.38), *IL2RA* (rs2476491, p=1.90 ⨯ 10^−8^, OR=1.22) and *AIRE* (rs74203920, p=2.33 ⨯ 10^−9^, OR=1.83) genes, thus providing the first robust associations between pernicious anemia and genetic risk factors. We further mapped the susceptibility in the HLA region to the HLA-DR15 haplotype. Analysis of associated diagnoses and disease trajectories confirm the association between pernicious anemia and thyroid issues, vitiligo, gastritis, stomach cancer, osteoporosis and other diagnoses.

## Main text

B12 deficiency anemia due to intrinsic factor deficiency, also known as pernicious anemia, is characterized by impaired B12 uptake caused by lack of intrinsic factor, a substance produced by epithelial cells of the stomach lining. Intrinsic factor normally binds B12 and facilitates absorption in the intestinal tract. Pernicious anemia is often caused by autoimmune damage to the stomach lining (autoimmune gastritis) in which case the gastric epithelial lining is damaged or destroyed^1^. The prevalence of pernicious anemia is around 0.1% in populations of European ancestry; however, it is more common in older people (∼2% in >60-year-olds), and believed to be less prevalent in Asian populations^2^. Symptoms of pernicious anemia range from fatigue to megaloblastic anemia and neurological abnormalities (peripheral numbness, paresthesia, ataxia) in more serious cases^3^.

Pernicious anemia is a complex disease with familial clustering, with a clear autoimmune basis and higher incidence of other autoimmune diseases, such as autoimmune thyroid conditions^4^, vitiligo^5^, type 1 diabetes^6^ in both patients with pernicious anemia and their relatives^7^. Although studies focusing on HLA serotypes have been conducted for pernicious anemia, the results have been conflicting^7^ and currently there is no clear consensus on the HLA alleles or other genetic risk factors predisposing to pernicious anemia.

We conducted a genome-wide association study (GWAS) of vitamin B12 deficiency due to lack of intrinsic factor to evaluate the contribution of genetic variation to the etiology of this disease in a combined dataset of 2,166 cases and 659,516 controls from three large population based biobanks - Estonian Biobank (EstBB)^8^, UK Biobank (UKBB)^9^, and FinnGen study. In the EstBB, individuals with pernicious anemia were identified using the ICD-10 code D51.0, resulting in 378 cases and 138,207 controls for analysis. Association testing was carried out with SAIGE 0.38 software^10^, adjusting for sex, year of birth and 10 PCs. Analysis in the EstBB was carried out under ethical approval 1.1-12/624 from the Estonian Committee on Bioethics and Human Research and data release N05 from the EstBB. GWAS summary statistics for the UKBB analysis including White British participants were downloaded from the UKBB PheWeb (http://pheweb.sph.umich.edu/SAIGE-UKB/about). Similarly, cases had been identified using the ICD-10 code D51.0 (754 cases, 390026 controls) and SAIGE had been used for association testing, adjusting for sex, birth year and the first 4 PCs. Summary statistics for FinnGen study were obtained from the publicly available R3 release PheWeb (https://www.finngen.fi/en/access_results). In the FinnGen study, we used the endpoint ‘Vitamin B12 deficiency anemia’, which included all the subcodes in the ICD10 D51 diagnosis group (1,034 cases, 131,283 controls). Similarly to other cohorts, SAIGE had been used for association testing, adjusting for sex, age, 10 PCs and genotyping batch. For meta-analysis, we used fixed-effects meta-analysis implemented in GWAMA^11^. Additional details available in **Methods** section.

In our GWAS meta-analysis we identify five genome-wide significant (p<5 × 10^−8^) associations (Table 1, Figure 1, Supplementary Figure 1) on 1p13.2 (lead signal rs6679677, p=1.91 × 10^−24^), 2p16.1 (rs12616502, p=3.14 × 10^−8^), 6p21.32 (rs28414666, p=1.40 × 10^−16^), 10p15.1 (rs2476491, p=1.90 × 10^−8^), and 21q22.3 (rs74203920, p=2.33 × 10^−9^), with similar effect estimates (effect heterogeneity was measured using Cochran’s test, p-values ranging from 0.05 to 0.83) in all analyzed cohorts (Figure 1), except for the lead variant on chromosome 2, which is absent in the Finnish data.

**Table 1.**
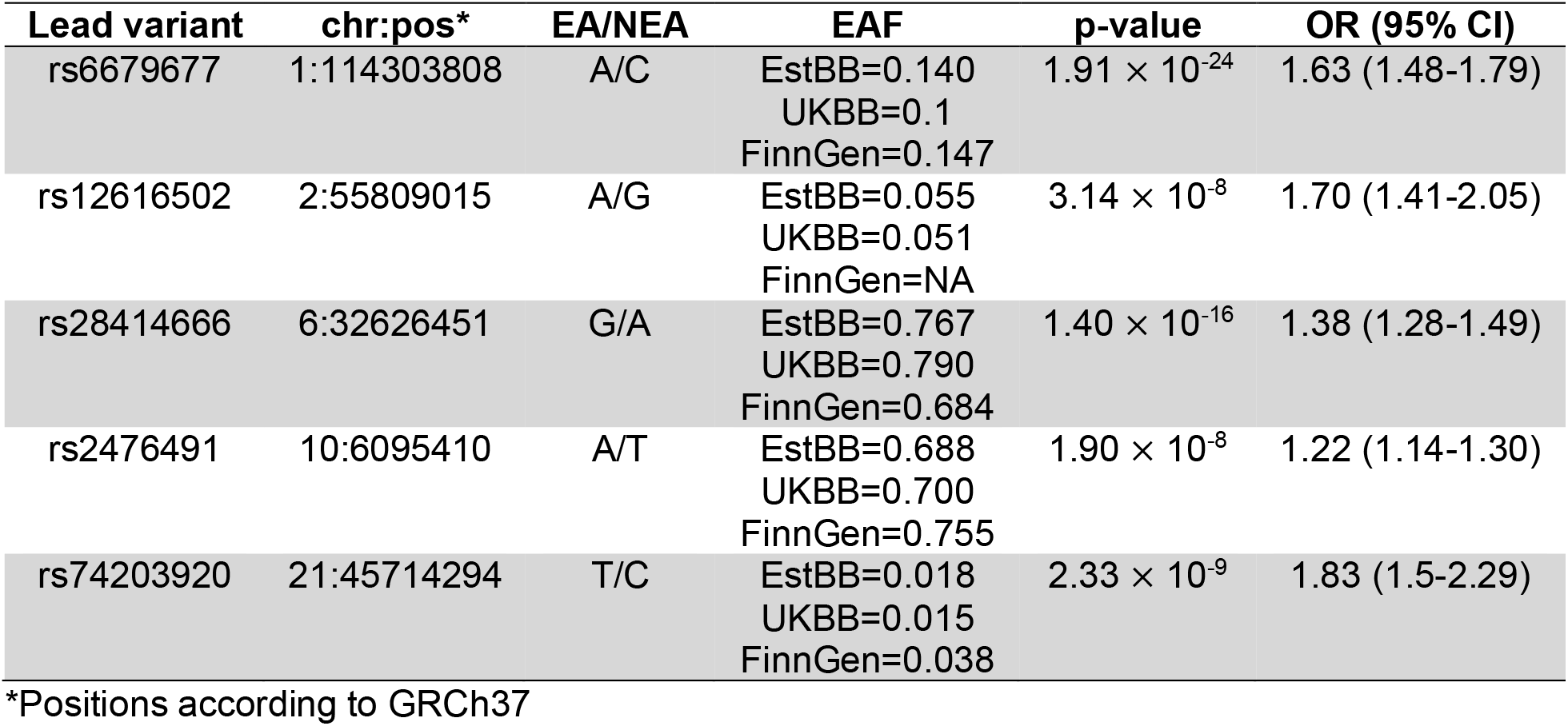
Summary of GWAS meta-analysis results for pernicious anemia.

| Lead variant | chr:pos* | EA/NEA | EAF | p-value | OR (95% CI) |
| --- | --- | --- | --- | --- | --- |
| rs6679677 | 1:114303808 | A/C | EstBB=0.140<br>UKBB=0.1<br>FinnGen=0.147 | $1.91 \times 10^{-24}$ | 1.63 (1.48-1.79) |
| rs12616502 | 2:55809015 | A/G | EstBB=0.055<br>UKBB=0.051<br>FinnGen=NA | $3.14 \times 10^{-8}$ | 1.70 (1.41-2.05) |
| rs28414666 | 6:32626451 | G/A | EstBB=0.767<br>UKBB=0.790<br>FinnGen=0.684 | $1.40 \times 10^{-16}$ | 1.38 (1.28-1.49) |
| rs2476491 | 10:6095410 | A/T | EstBB=0.688<br>UKBB=0.700<br>FinnGen=0.755 | $1.90 \times 10^{-8}$ | 1.22 (1.14-1.30) |
| rs74203920 | 21:45714294 | T/C | EstBB=0.018<br>UKBB=0.015<br>FinnGen=0.038 | $2.33 \times 10^{-9}$ | 1.83 (1.5-2.29) |

\*Positions according to GRCh37

**Figure 1.**
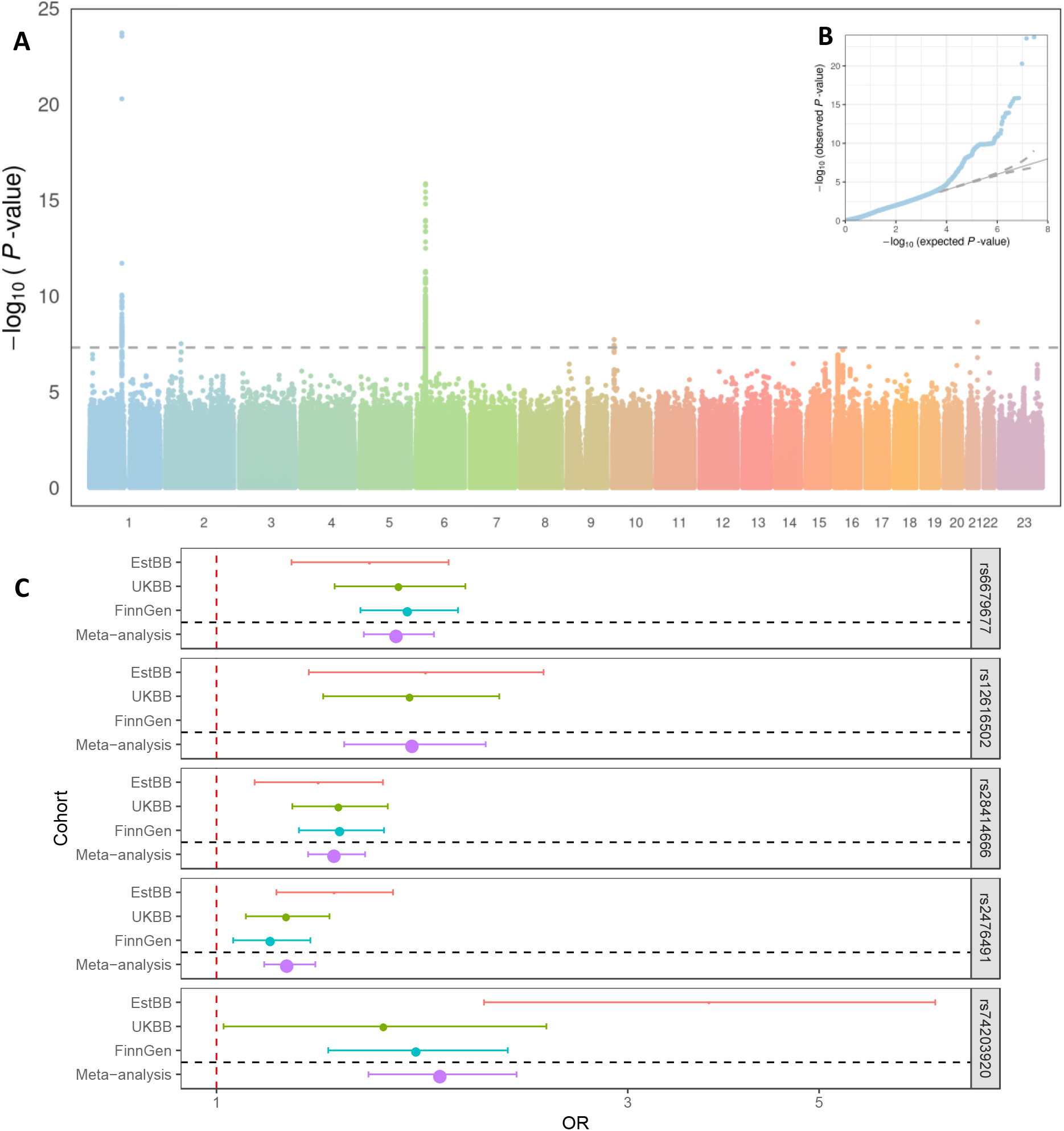
Results of the pernicious anemia GWAS meta-analysis. A) Manhattan and B) QQ plot; C) Forest plot of effect estimates for lead variants associated with pernicious anemia. The odds ratios (dots) and 95% confidence intervals (error bars) are shown for all included cohorts and meta-analysis. The size of the dot is proportional to the effective sample size, (calculated as 4/((1/N_cases)+(1/N_controls)). Pernicious anemia is defined as ICD10 code D51.0 in EstBB and UKBB and as D51 (vitamin B12 deficiency anemia) in FinnGen.

The lead SNP on chr1, rs6679677, is in high LD (r^2^=0.96) with a non-synonymous variant in exon 12 of the *PTPN22* gene (rs2476601, p=2.82 × 10^−24^). *PTPN22* is a known immune regulator gene and this particular variant has been associated with several autoimmune diseases, including rheumatoid arthritis, Crohn’s disease, systemic lupus erythematosus, vitiligo, autoimmune thyroid conditions, type 1 diabetes and others (Supplementary Table 1 and 3, Supplementary Figure 2).

On chromosome 2, the lead signal rs12616502 is in high LD with exonic non-synonymous variants in *CCDC104* (r^2^=0.94, rs1045920) and *PNPT1* (r^2^=0.94, rs7594497), and this region has been previously associated with both vitiligo, hypothyroidism and myelodysplastic syndrome (Supplementary Tables 1 and 3, Supplementary Figure 2). Colocalization analysis demonstrated that pernicious anemia has a common causal variant with the expression of *PTPN1* in thyroid tissue in GTEx v8 dataset (Figure 2, Supplementary Table 2). *PTPN1* is further supported as a candidate causal gene in this locus by data from mouse knockouts, as *Pnpt1tm1a(KOMP)Wtsi/Pnpt1+* mice exhibit increased mean corpuscular volume (MCV) together with increased mean corpuscular hemoglobin (MCH)^12^. Increased red blood cell MCV is a common feature in macrocytic anemias, both megaloblastic (caused by B12 deficiency and pernicious anemia) and nonmegaloblastic (caused by diseases such as myelodysplastic syndrome and hypothyroidism)^13^.

**Figure 2.**
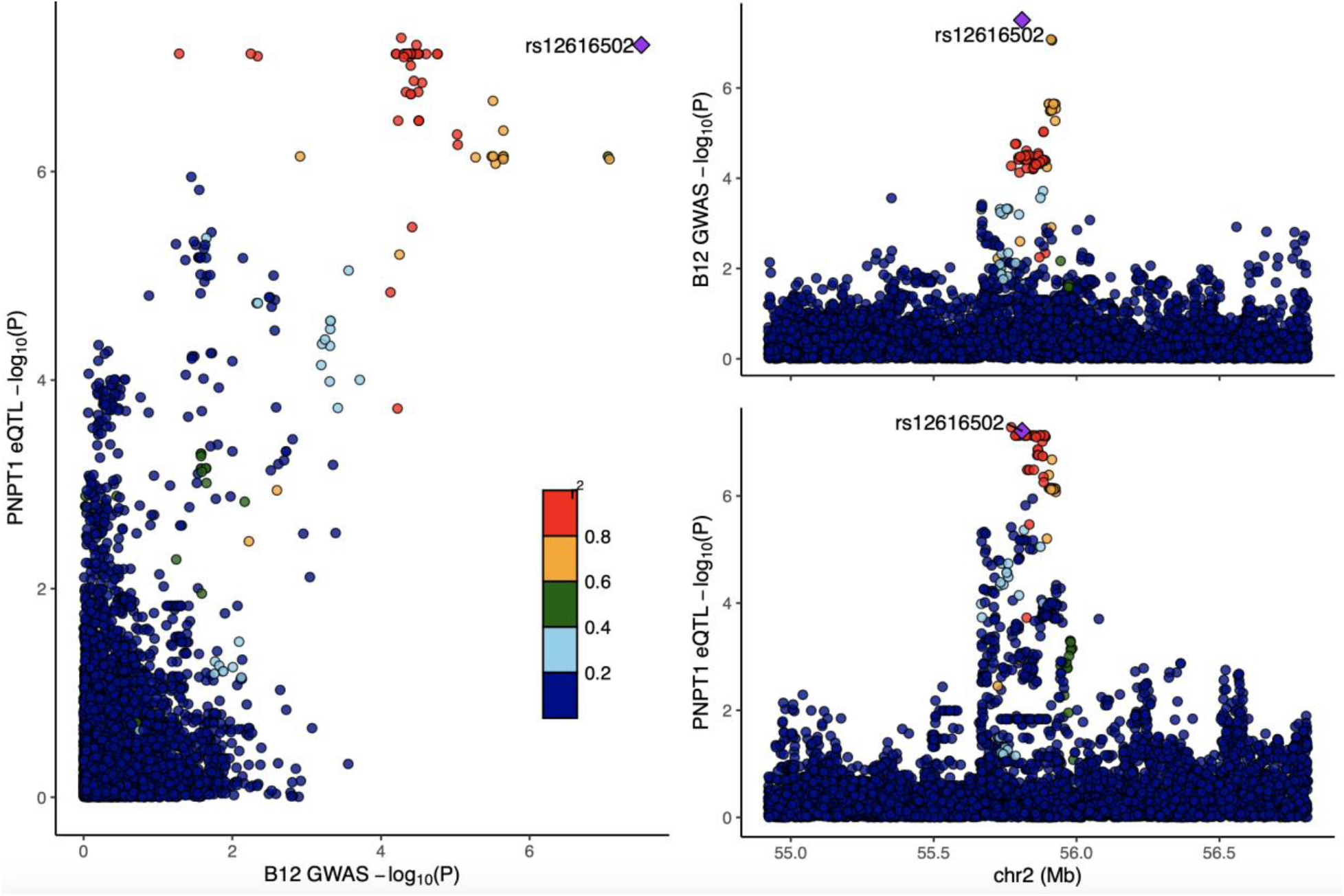
GWAS signature for pernicious anemia co-localizes with the eQTL signal for *PNPT1* in thyroid tissue in GTEx v8 dataset (posterior probability for shared causal variant PP4=0.87). Left – comparison on -log10 p-values from pernicious anemia GWAS-meta-analysis (x-axis) and eQTL analysis(y-axis). Right – locus plots for the *PNPT1* locus from GWAS meta-analysis (top) and eQTL analysis (bottom). For all plots, LD is colored with respect to the GWAS lead signal rs12616502 (labeled).

The third signal on chromosome 6 is located in the HLA region, a common hub for autoimmune condition associations, downstream *HLA-DQB1*. To further clarify the association signal in the HLA region, we used data on a total of 249 imputed HLA alleles at four-digit level available for 183 cases and 67,505 controls in the EstBB. Three tested alleles passed the Bonferroni corrected threshold of association (0.05/249=2 × 10^−4^), including HLA-DRB1*15:01 (p=1.3 × 10^−6^), HLA-DQB1*06:02 (p=1.3 × 10^−6^) and HLA-DQA1*01:02 (p=2.8 × 10^−5^). The DRB1*15:01-DQB1*06:02-DQA1*01:02 combination forms the HLA-DR15 haplotype, which is a reported risk factor for multiple sclerosis^14^. HLA-DR15 belongs under the HLA-DR2 group, which has been associated with pernicious anemia in a 1981 study^15^.

On chromosome 10, the sentinel variant rs2476491 is intronic to *IL2RA. IL2RA* encodes the interleukin-2 receptor alpha chain, thus being involved in regulating regulatory T-cells and immune tolerance, as regulatory T cells suppress autoreactive T-cells. Accordingly, this locus has previously been associated with multiple sclerosis, juvenile idiopathic arthritis, vitiligo and hypothyroidism (Supplementary Table 1).

Finally, the association on chromosome 21, rs74203920, is a missense variant in the *AIRE* gene, a known autoimmune regulator. Interestingly, mutations in *AIRE* are a known cause of autoimmune polyendocrinopathy syndrome type 1 (APS-1), which is a rare autosomal recessive syndrome, that sometimes includes pernicious anemia among other components^16^.

We used the individual level data in EstBB to evaluate the association between pernicious anemia and other diseases (defined by ICD-10 codes). According to our analysis, individuals with pernicious anemia have more diagnoses of other anemias and vitamin deficiencies (Figure 3, Supplementary Table 4), thyroid problems (thyroiditis and hypothyroidism), gastrointestinal tract diagnoses (gastritis, malignant neoplasm of stomach, intestinal malabsorption, irritable bowel syndrome), but also of vitiligo, dermatitis, osteoporosis and spontaneous abortion. Evaluation of disease timeline shows that while there is considerable individual variation, on average, thyroid issues (ICD10 E03 other hypothyroidism and E06 thyroiditis, Figure 3), intestinal problems (K12 stomatitis, K29 gastritis and K58 irritable bowel syndrome), spontaneous abortion (O03), depression (F32) and vitiligo (L80) are diagnosed before pernicious anemia, while nutritional deficiencies are diagnosed after (D52 folate deficiency, E53 deficiency of other B vitamins, E55 vitamin D deficiency, E61 deficiency of other nutrient elements). Majority of these diagnoses reflect the etiology of pernicious anemia (gastritis) or symptoms (skin problems, syncope and collapse, depression, stomatitis), known comorbidities (vitiligo, thyroid issues^4,5^), or diseases where pernicious anemia is a known risk factor (such as osteoporosis^17^ and stomach cancer^18^). The association with spontaneous abortion is interesting, as although there is some evidence B12 deficiency and pernicious anemia could cause recurrent miscarriage^19–21^, the data is scarce and the link with spontaneous miscarriage has not been explored in depth.

**Figure 3.**
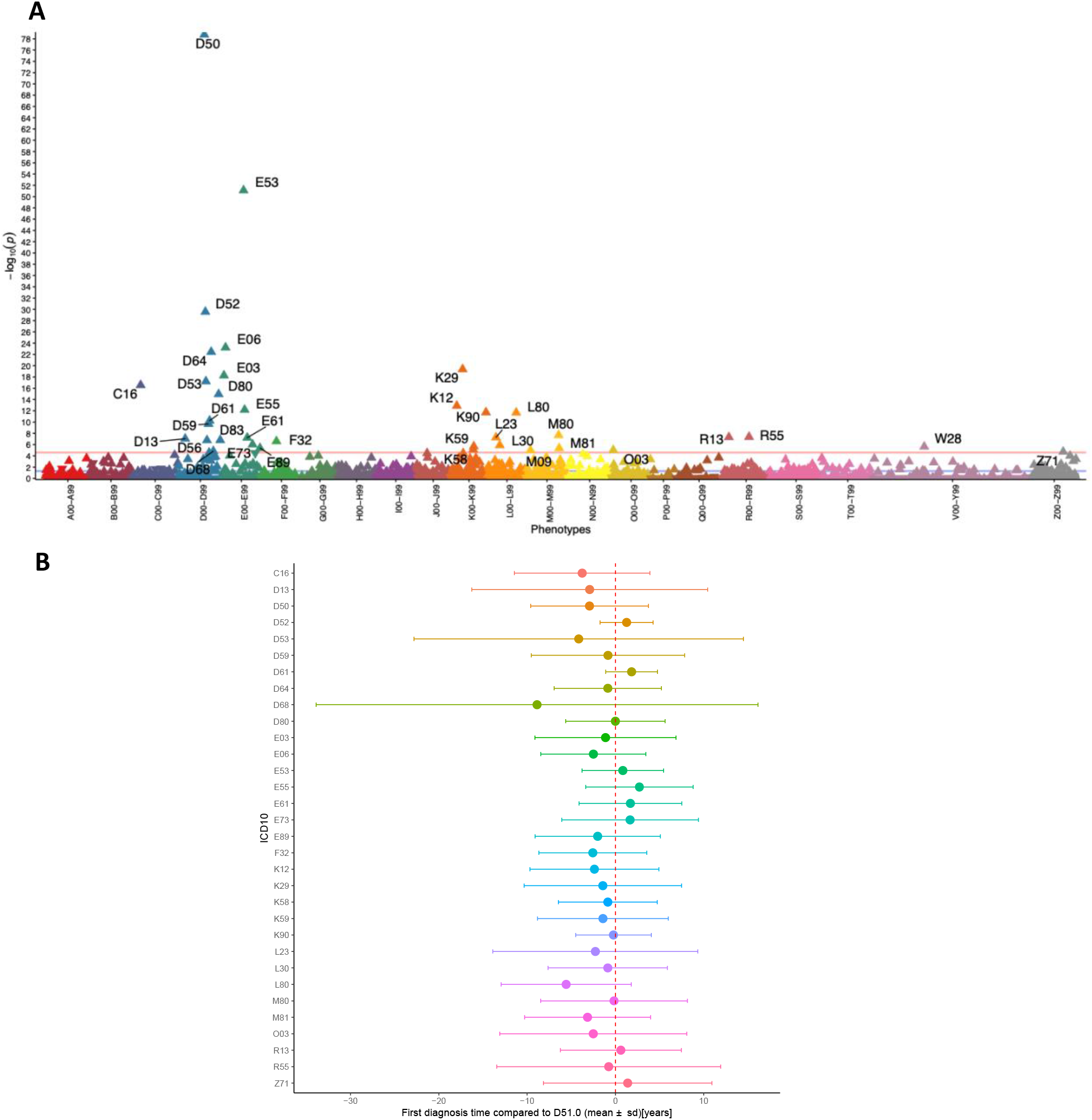
Association plot for ICD10 codes associated with pernicious anemia diagnosis in Estonian Biobank. A) Each triangle in the plot represents one ICD10 main code and the direction of the triangle represents direction of effect – upward-pointing triangles show increased prevalence of diagnosis code in pernicious anemia cases. Red line – Bonferroni-corrected significance level (2.5 × 10^−5^). B) Time at first diagnosis of associated diagnosis code compared to time of pernicious anemia (D51.0) diagnosis. Plot shows mean difference in years, error bars correspond to standard deviation. Red dashed vertical line represents D51.0 diagnosis.

This GWAS meta-analysis focused on individuals with pernicious anemia identified from population-based biobanks, thus more detailed analysis on the effect these genetic risk factors have on disease severity or subphenotypes is unfortunately not possible. Although the used summary statistics for FinnGen cohort are for a broader vitamin B12 deficiency phenotype definition compared toother two cohorts, we see no significant differences in effect estimates for the reported lead variants. The analyses included participants of European ancestry, however, analyses in other populations are warranted, since the prevalence of pernicious anemia differs depending on racial background, being more common in people with African or European ancestry, especially from Scandinavia and UK^2^. The analysis of associated phenotypes could potentially provide clinically useful insights into pernicious anemia disease trajectories and offer information for patient management; however, at the moment it only includes participants of the EstBB, with limited follow-up time, therefore further studies are needed.

In summary, our analysis of 2,166 cases and 659,516 controls identifies robust risk loci for pernicious anemia in or near candidate genes with a known role in autoimmune conditions (*PTPN22, HLA, IL2RA, AIRE*). We further narrow the signal in the HLA region to the HLA-DR15 haplotype and propose *PNPT1* as the most likely causal gene in the 2p16.1 locus. The associations between the identified loci and other autoimmune conditions, such as type 1 diabetes, vitiligo, and autoimmune thyroid conditions help to clarify the link between pernicious anemia and its common comorbidities. Analysis of associated diagnoses and disease trajectories confirm the association between pernicious anemia and thyroid issues, vitiligo, gastritis, stomach cancer, osteoporosis and other diagnoses, but also between pernicious anemia and spontaneous abortion.

## Methods

### Cohorts

#### Estonian Biobank

The Estonian Biobank is a population-based biobank with over 200,000 participants. The 150K data freeze was used for the analyses described in this paper. All biobank participants have signed a broad informed consent form. Individuals with pernicious anemia were identified using the ICD-10 code D51.0, and all biobank participants who did not have this diagnosis were considered as controls. Information on ICD codes is obtained via regular linking with the national Health Insurance Fund and other relevant databases^8^.

All EstBB participants have been genotyped at the Core Genotyping Lab of the Institute of Genomics, University of Tartu, using Illumina GSAv1.0, GSAv2.0, and GSAv2.0_EST arrays. Samples were genotyped and PLINK format files were created using Illumina GenomeStudio v2.0.4. Individuals were excluded from the analysis if their call-rate was < 95% or if sex defined based on heterozygosity of X chromosome did not match sex in phenotype data. Before imputation, variants were filtered by call-rate < 95%, HWE p-value < 1e-4 (autosomal variants only), and minor allele frequency < 1%. Variant positions were updated to b37 and all variants were changed to be from TOP strand using GSAMD-24v1-0_20011747_A1-b37.strand.RefAlt.zip files from https://www.well.ox.ac.uk/~wrayner/strand/ webpage. Prephasing was done using Eagle v2.3 software^22^ (number of conditioning haplotypes Eagle2 uses when phasing each sample was set to: --Kpbwt=20000) and imputation was done using Beagle v.28Sep18.793^23^ with effective population size ne=20,000. Population specific imputation reference of 2297 WGS samples was used.

Association analysis was carried out using SAIGE (v0.38)^10^ software implementing mixed logistic regression model with LOCO option, using sex, year of birth and 10 PCs as covariates in step I.

#### UK Biobank

The UK Biobank (UKBB) is a prospective cohort of 502,637 individuals aged 37-73 recruited in 2006-2010 from across the UK, who completed detailed questionnaires regarding socio-demographic and lifestyle characteristics and their medical history and had a clinical assessment. Additional information about medical conditions (both existing at baseline and occurring during follow-up) has been obtained through linking with hospital admission and mortality data. Full details of the study have been reported in Sudlow et al^9^. Publicly available GWAS summary statistics downloaded from the UKBB PheWeb [http://pheweb.sph.umich.edu/SAIGE-UKB/about] were used for the analysis. Briefly, the PheWeb includes GWAS summary statistics for ICD code-based traits extracted from electronic health records. Phenotypes have been classified into 1,403 broad PheWAS codes, including pernicious anemia (PheCode 281.11), defined using the ICD-10 code D51.0 and excluding other anemias under the PheCodes 280-285.99. Genetic analyses have been carried out using SAIGE^10^.

#### FinnGen

FinnGen is a public-private partnership project combining data from Finnish biobanks and electronic health records from different registries. After a one-year embargo, the FinnGenv summary stats are available for download. In this study, we used the results from the FinnGen release R3, which includes data from 135,638 individuals and more than 1,800 disease endpoints. FinnGen individuals have been genotyped with Illumina and Affymetrix arrays and imputed to the population-specific SISu v3 importation reference panel. Genetic association testing has been carried out with SAIGE^10^. The FinnGen disease endpoint “Vitamin B12 deficiency anemia” included all individuals with the ICD10 D51 diagnosis as cases. For more information on genotype data, disease endpoints and GWAS analyses, please see https://finngen.gitbook.io/documentation/.

### GWAS meta-analysis

We extracted all genetic variants with a rs-number from the summary statistics of the three participating cohorts and conducted an inverse of variance weighted fixed-effects meta-analysis without genomic control using GWAMA^11^. A total of 30,907,385 variants were included in the meta-analysis. Genome-wide significance was set to p < 5 × 10^−8^.

### HLA allele imputation in the EstBB

Imputation of HLA alleles from SNP data was carried out at the Broad Institute using the SNP2HLA tool^24^.

### Colocalization

We conducted colocalization analyses to detect shared causal variants between pernicious anemia and gene expression using COLOC (v.3.2.1) R package^25^ and GWAS meta-analysis summary statistics. We set the prior probabilities to p_1_=1 × 10^−4^, p_2_=1 × 10^−4^, p_12_=5 × 10^−6^ as suggested by Wallace (2020)^26^ and used the COLOC version which takes regression coefficients and their variance into account.

In the analysis we compared our significant GWAS loci to all eQTL Catalogue (https://www.ebi.ac.uk/eqtl/) RNA-seq datasets (excluding Lepik et al. 2017^27^ due to sample overlap) containing QTLs for gene expression, exon expression, transcript usage and txrevise event usage; eQTL Catalogue microarray datasets containing QTLs for gene expression (excluding Kasela et al 2017^28^); and GTEx v8 datasets containing QTLs for gene expression (see Methods: https://www.ebi.ac.uk/eqtl/Methods/).

We lifted the GWAS summary statistics over to hg38 build to match the eQTL Catalogue. For each genome-wide significant (p<5 × 10^−8^) GWAS locus we extracted the 1Mbp radius of its top hit from QTL datasets and ran the colocalization analysis for those eQTL Catalogue traits that had at least one cis-QTL within this region with p< 1 × 10^−6^. We considered two signals to colocalize if the posterior probability for a shared causal variant was 0.8 or higher. All results with a PP4 > 0.8 can be found in Supplementary Table 2.

Results were visualized with the *LocusCompareR* library^30^.

### Look-up of phenome-wide associations in GWAS catalog and with PhenoScanner v2

FUMA v1.3.6a^29^ was used to compare the genome-wide significant lead signals and markers in high LD with these markers against the results in the GWAS catalogue. The results of this look-up are presented in Supplementary Table 1.

PhenoScanner v2^31,32^ was used for look-up of phenotype associations for the GWAS lead variants in previous GWAS studies. PhenoScanner query was done using the rsid-s of GWAS lead variants and the *phenoscanner* R package (https://github.com/phenoscanner/phenoscanner). Query results were filtered to keep one association per variant per trait, keeping studies from newer or larger studies. Descriptions of Experimental Factor Ontology (EFO) terms and classification of EFO broad categories were obtained from the GWAS Catalog. Missing categories were added by manually searching the EMBL-EBI EFO webpage (www.ebi.ac.uk/efo/). For visualization of PhenoScanner results, parent categories with fewer results were grouped into larger categories and a heatmap was created using the *pheatmap* library in R 3.6.1. and a modified script from (https://github.com/LappalainenLab/spiromics-covid19-eqtl/blob/master/eqtl/summary_phenoscanner_lookup.Rmd). The results of this look-up are presented in Supplementary Table 3.

### Mouse phenotypes

We used the Mouse Genome Database^12^ (http://www.informatics.jax.org) to evaluate the *PNPT1* effect on phenotype in mouse models.

### Analysis of associated phenotypes in EstBB

Using the individual level data in the EstBB, we conducted an analysis to find ICD10 diagnosis codes associated with the D51.0 diagnosis. We tested the association between pernicious anemia status (defined as ICD10 D51.0) and other ICD10 codes using logistic regression and adjusting for sex, age and 10 PCs. Bonferroni correction was applied to select statistically significant associations (Number of tested ICD main codes – 1,944, corrected p-value threshold – 2.5 × 10^− 5^). Results were visualised using the *PheWas* library (https://github.com/PheWAS/PheWAS). All analyses were carried out in R 3.6.1. Diagnosis timeline of pernicious anemia and significantly associated diagnoses was evaluated by calculating the time interval (in years) between the first date of pernicious anemia diagnosis (D51.0) and first date of associated disease for each individual. Results were visualized by plotting the means and standard deviations of calculated intervals using R 3.6.1. The results of this analysis are presented in Supplementary Table 4.

## Data Availability

Used UKBB and FinnGen summary statistics can be browsed and downloaded from UKBB PheWeb and FinnGen PheWeb, respectively. Full meta-analysis summary statistics will be made available upon publication. The analyses in this manuscript also included data from the Mouse Genome Database and the GTEx Portal.

http://pheweb.sph.umich.edu/SAIGE-UKB/about

http://r3.finngen.fi

http://www.informatics.jax.org

https://www.ebi.ac.uk/eqtl/

## Data availability

Used UKBB and FinnGen summary statistics can be browsed and downloaded from UKBB PheWeb and FinnGen PheWeb, respectively. Full meta-analysis summary statistics will be made available upon publication.

## Web resources

UKBB PheWeb: http://pheweb.sph.umich.edu/SAIGE-UKB/about; FinnGen Freeze 3 PheWeb: http://r3.finngen.fi; Mouse Genome Database: http://www.informatics.jax.org; GTEx Portal: https://gtexportal.org/home/; eQTL Catalogue: https://www.ebi.ac.uk/eqtl/

## Acknowledgements

TL is supported by the Estonian Research Council grant MOBTP155. ML and RM are supported by the Estonian Research Council grant PRG687. Computations were performed in the High Performance Computing Center, University of Tartu. We want to acknowledge the participants and investigators of the FinnGen and UKBB studies. The Genotype-Tissue Expression (GTEx) Project was supported by the Common Fund of the Office of the Director of the National Institutes of Health, and by NCI, NHGRI, NHLBI, NIDA, NIMH, and NINDS. The data used for the analyses described in this manuscript were obtained from the GTEx Portal on 10/01/20.

