## Supplementary Figure 1 for "Genome-wide association study identifies five risk loci for pernicious anemia and implicates the role of HLA-DR15 haplotype"

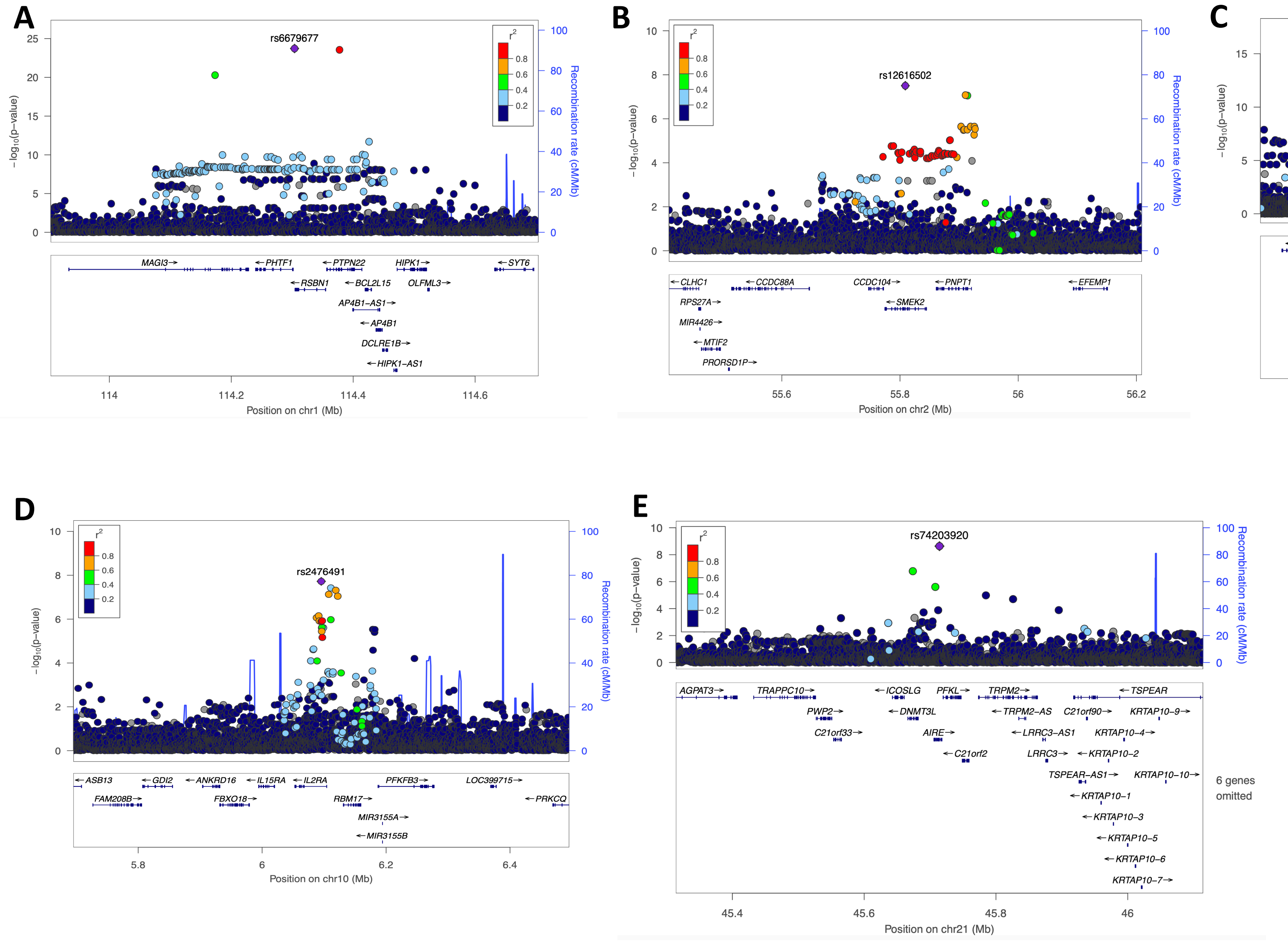


**Supplementary Figure 1.** Regional plots of five association signals from pernicious anemia GWAS meta-analysis. A) rs6679677 on chr1; B) rs12616502 on chr2; C) rs28414666 on chr6; D) rs2476491 on chr10; and E) rs74203920 on chr21
