## Supplementary figures and images for "Genome-wide association study identifies five risk loci for pernicious anemia and implicates the role of HLA-DR15 haplotype"

### Supplementary Figure 2

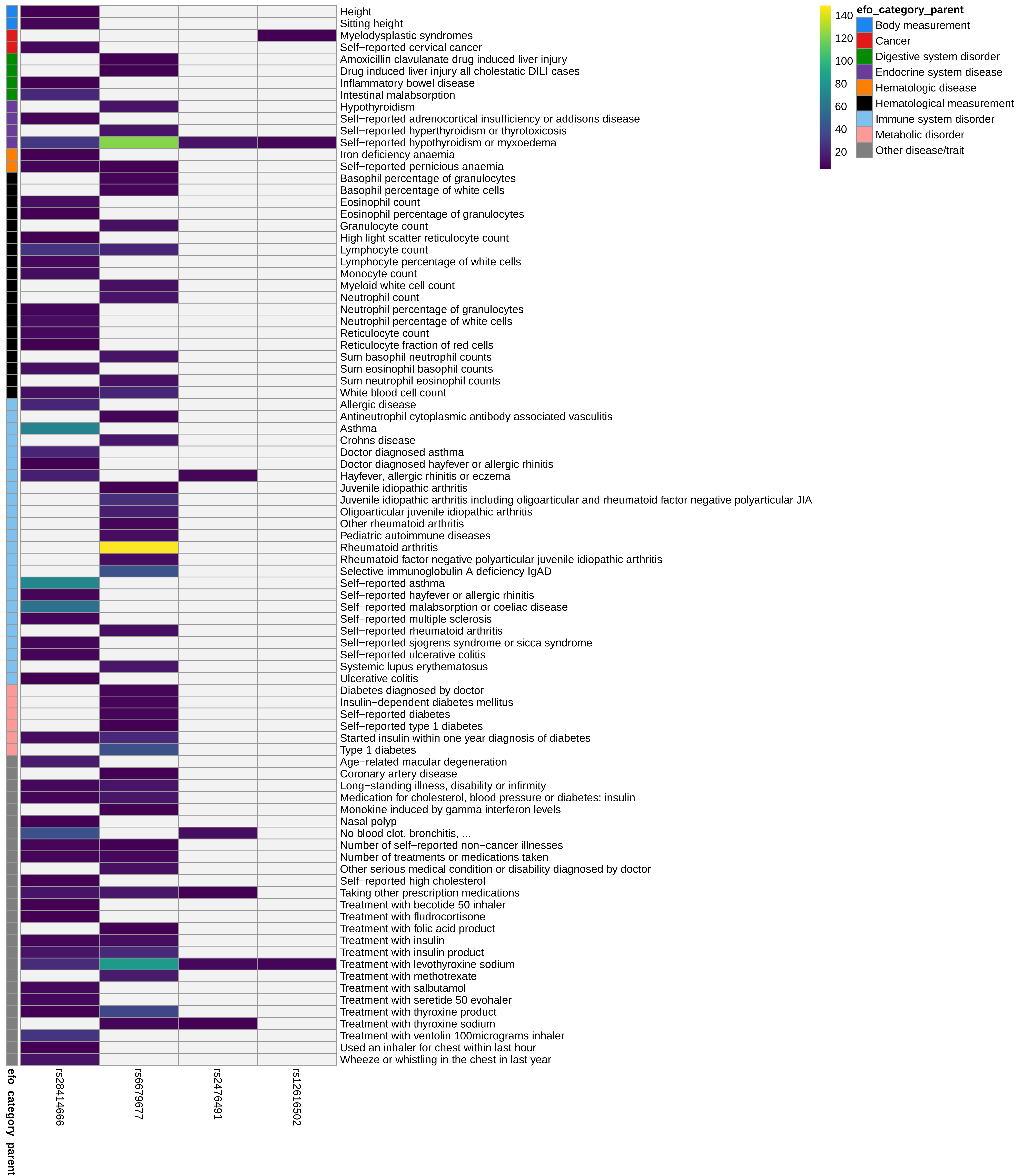
